# Effectiveness of influenza vaccination against SARS-CoV-2 infection among healthcare workers in Qatar

**DOI:** 10.1101/2022.05.09.22274802

**Authors:** Elias Tayar, Sami Abdeen, Muna Abed Alah, Hiam Chemaitelly, Iheb Bougmiza, Houssein H. Ayoub, Anvar Hassan Kaleeckal, Ali Nizar Latif, Riyazuddin Mohammad Shaik, Hamad Eid Al-Romaihi, Mohamed H. Al-Thani, Roberto Bertollini, Laith J. Abu-Raddad, Abdullatif Al-Khal

**Affiliations:** Community Medicine Department, Hamad Medical Corporation, Doha, Qatar; Infectious Disease Epidemiology Group, Weill Cornell Medicine-Qatar, Cornell University, Doha, Qatar; World Health Organization Collaborating Centre for Disease Epidemiology Analytics on HIV/AIDS, Sexually Transmitted Infections, and Viral Hepatitis, Weill Cornell Medicine–Qatar, Cornell University, Qatar Foundation – Education City, Doha, Qatar; Department of Population Health Sciences, Weill Cornell Medicine, Cornell University, New York, New York, USA; Community Medicine Department, Primary Health Care Corporation, Doha, Qatar; Community Medicine Department, College of Medicine, Sousse University, Tunisia; Mathematics Program, Department of Mathematics, Statistics, and Physics, College of Arts and Sciences, Qatar University, Doha, Qatar; Business Intelligence and Operational Performance Unit, Hamad Medical Corporation, Doha, Qatar; Ministry of Public Health, Doha, Qatar; Department of Public Health, College of Health Sciences, QU Health, Qatar University, Doha, Qatar

**Author notes:** Correspondence to Professor Laith J. Abu-Raddad,. These authors contributed equally (Elias Tayar, Sami Abdeen, Muna Abed Alah).

**Keywords:** influenza, COVID-19, vaccine, test-negative, case-control, immunity, epidemiology

## Abstract

A number of studies reported that influenza vaccination is associated with lower risk of severe acute respiratory syndrome coronavirus 2 (SARS-CoV-2) infection and/or coronavirus disease 2019 (COVID-19) morbidity and mortality. We conducted a matched, test-negative, case-control study to estimate effectiveness of influenza vaccination, using Abbott’s quadrivalent Influvac Tetra vaccine, against SARS-CoV-2 infection and against severe COVID-19. The study was implemented on a population of 30,774 healthcare workers (HCWs) in Qatar during the 2020 annual influenza vaccination campaign, between September 17, 2020 and December 31, 2020, before introduction of COVID-19 vaccination. The median age in the matched samples was 36 years (interquartile range (IQR), 32-44) for cases and 35 years (IQR, 32-42) for controls. The median duration between influenza vaccination and the PCR test was 43 days (IQR, 29-62). The estimated effectiveness of influenza vaccination against SARS-CoV-2 infection >14 days after receiving the vaccine was 29.7% (95% CI: 5.5-47.7%). The estimated effectiveness of influenza vaccination against any severe, critical, or fatal COVID-19 was 88.9% (95% CI: 4.1-98.7%). Sensitivity analyses confirmed main analysis results. Recent influenza vaccination is associated with an appreciable reduction in the risk of SARS-CoV-2 infection and COVID-19 severity.

## Introduction

Influenza vaccination protects against influenza infection and reduces the morbidity and mortality of seasonal influenza^1^. This vaccination is strongly recommended for high-risk groups such as the elderly and healthcare workers (HCWs)^2^. A number of studies have reported that influenza vaccination is also associated with lower risk of severe acute respiratory syndrome coronavirus 2 (SARS-CoV-2) infection and/or coronavirus disease 2019 (COVID-19) morbidity and mortality^3-12^. However, reported apparent protection against SARS-CoV-2 infection and severe COVID-19 could have arisen due to bias and may not have reflected a genuine biological effect. In particular, this protection may have been due to the healthy user effect^13^, whereby health-aware persons are more likely to receive an influenza vaccine, and simultaneously, to practice health behaviors that reduce their risk of acquiring the infection^3,5^.

Against this background, we assessed the effectiveness of influenza vaccination, using Abbott’s quadrivalent Influvac Tetra vaccine, against SARS-CoV-2 infection and against any severe (acute-care hospitalization),^14^ critical (intensive-care-unit hospitalization),^14^ or fatal^15^ COVID-19 among HCWs in Qatar during the 2020 annual influenza vaccination campaign, and before introduction of COVID-19 vaccination. Since variation in health behaviors among HCWs is presumably less than that among the general population, conducting this study among HCWs allowed us to minimize the influence of the healthy user effect on effectiveness estimates.

## Methods

### Study design

This study was conducted among HCWs at Hamad Medical Corporation, the principal provider of public healthcare services in Qatar and the nationally designated entity for COVID-19-related healthcare needs. The study analyzed the national, federated databases for COVID-19 laboratory testing, vaccination, hospitalization, and death, retrieved from the integrated nationwide digital-health information platform. These databases include all SARS-CoV-2-related data and associated demographic information, with no missing information, since the pandemic onset, documenting all polymerase chain reaction (PCR) tests. Influenza vaccination data were retrieved from Hamad Medical Corporation’s influenza vaccination database. Further descriptions of these databases have been reported previously^16-22^.

Every PCR test conducted in Qatar is classified on the basis of symptoms and the reason for testing (clinical symptoms, contact tracing, surveys or random testing campaigns, individual requests, routine healthcare testing, pre-travel, screening at ports of entry, or other). Qatar has unusually young, diverse demographics, in that only 9% of its residents are ≥50 years of age, and 89% are expatriates from over 150 countries^16^. This diversity extends to the HCW population.

Effectiveness of influenza vaccination against SARS-CoV-2 infection was assessed using the test-negative, case-control study design, a preferred design for assessing vaccine effectiveness against influenza^23,24^. This design was also validated in Qatar’s population and has been repeatedly applied in previous studies in this country to assess COVID-19 vaccine effectiveness^17^,^19^,^20^,^25-28^.

All HCWs with a SARS-CoV-2 PCR test performed between September 17, 2020 and December 31, 2020 were eligible for inclusion in the study. This duration coincided with the national annual influenza vaccination campaign that typically starts in September or October, and continues for several months thereafter. Influenza vaccination is offered free of charge to all citizens and residents. During the 2020 influenza vaccination campaign, only Abbott’s quadrivalent Influvac Tetra vaccine was used. The study was concluded at the start of the mass COVID-19 vaccination campaign. During the study, SARS-CoV-2 incidence was due to the original SARS-CoV-2 virus, before introduction of SARS-CoV-2 variants of concern^29-32^. SARS-CoV-2 incidence was also relatively low during the study with no wave materializing during this time^33^.

To estimate effectiveness against SARS-CoV-2 infection, we exact-matched cases (HCWs with PCR-positive tests) and controls (HCWs with PCR-negative tests) identified during the study in a 1:5 ratio by sex, 10-year age groups, 10-nationality groups, reason for PCR testing, and bi-weekly PCR test date, to control for known differences in SARS-CoV-2 exposure risk in Qatar^16,33-36^. Matching by these factors was shown previously in studies of different epidemiologic designs to provide adequate control of differences in the risk of exposure to SARS-CoV-2 infection in Qatar^17,20,22,25,37^.

Only the first PCR-positive test for the cases and the first PCR-negative test for the controls during the study were included in the analysis. Controls included individuals with no record of a PCR-positive test during the study period. HCWs with a PCR test within 14 days after receiving the influenza vaccine and those who received COVID-19 vaccination were excluded. These inclusion and exclusion criteria were implemented to allow adequate time for build-up of immunity after vaccination^19^, and to minimize different types of potential bias, as informed by earlier analyses in the same population^17,25^. Every control that met the inclusion criteria and that could be matched to a case was included in the analysis.

Classification of COVID-19 case severity^14^, criticality^14^, and fatality^15^ followed World Health Organization guidelines, and assessments were made by trained medical personnel using individual chart reviews, as part of a national protocol applied to every hospitalized COVID-19 patient. Details of COVID-19 severity, criticality, and fatality classifications are found in Supplementary Section S1.

Every hospitalized COVID-19 patient underwent infection severity assessment every three days until discharge or death. We classified individuals who progressed to severe, critical, or fatal COVID-19 between the time of the PCR-positive test and the end of the study based on their worst disease outcome, starting with death^15^, followed by critical disease^14^, and then severe disease^14^.

### Laboratory methods

Details of laboratory methods for real-time reverse-transcription PCR (RT-qPCR) testing are found in Supplementary Section S2. All PCR testing was conducted at the Hamad Medical Corporation Central Laboratory or at Sidra Medicine Laboratory, following standardized protocols.

### Statistical analysis

Baseline characteristics of cases and controls were reported using descriptive statistics. Groups were compared using standardized mean differences (SMDs), with SMD close to 0.1 indicating adequate matching^38^. Conditional logistic regression factoring the matched design was performed to compare the odds of influenza vaccination in cases versus controls. This analytical approach, including matching by bi-weekly PCR test date, minimizes potential bias due to variation in epidemic phase^23,39^ and roll-out of vaccination during the study^23,39^. Interactions were not considered. The resulting estimates for the adjusted odds ratio and 95% confidence interval (CI) were then used to estimate influenza vaccination effectiveness and corresponding 95% CI using the equation^23,24^:

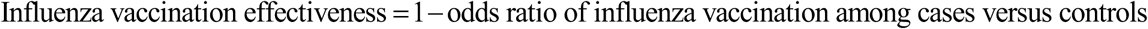

Effectiveness was estimated against SARS-CoV-2 infection, whether symptomatic or asymptomatic, and against any severe, critical, or fatal COVID-19. Sensitivity analyses were conducted to investigate the impact of adjusting for prior infection in the conditional logistic regression, and of modifying the study inclusion and exlusion criteria on the estimate of influenza vaccination effectiveness. The latter was informed by an established analysis plan to invistigate sources of potential bias^17,25^, and encomapssed exclusion of individuals with prior infection, exclusion of travel-related PCR testing, or inclusion of all PCR-positive and PCR-negative tests for cases and controls. Statistical analyses were conducted using STATA/SE version 17.0 (Stata Corporation, College Station, TX, USA).

### Oversight

Hamad Medical Corporation and Weill Cornell Medicine-Qatar Institutional Review Boards approved this retrospective study with a waiver of informed consent. The research was performed in accordance with relevant guidelines and regulations. The study was reported following the Strengthening the Reporting of Observational Studies in Epidemiology (STROBE) guidelines. The STROBE checklist can be found in Table S1.

## Results

Figure 1 shows the process of selecting the study population. Of 30,774 HCWs at Hamad Medical Corporation, 12,015 had one or more SARS-CoV-2 PCR tests between September 17, 2020 and December 31, 2020. Of these, 576 HCWs with a PCR-positive test and 10,033 with exlusively PCR-negative tests were egligible for inclusion in the study. Exact-matching on a 1:5 ratio yielded 518 cases matched to 2,058 controls.

**Figure 1.**
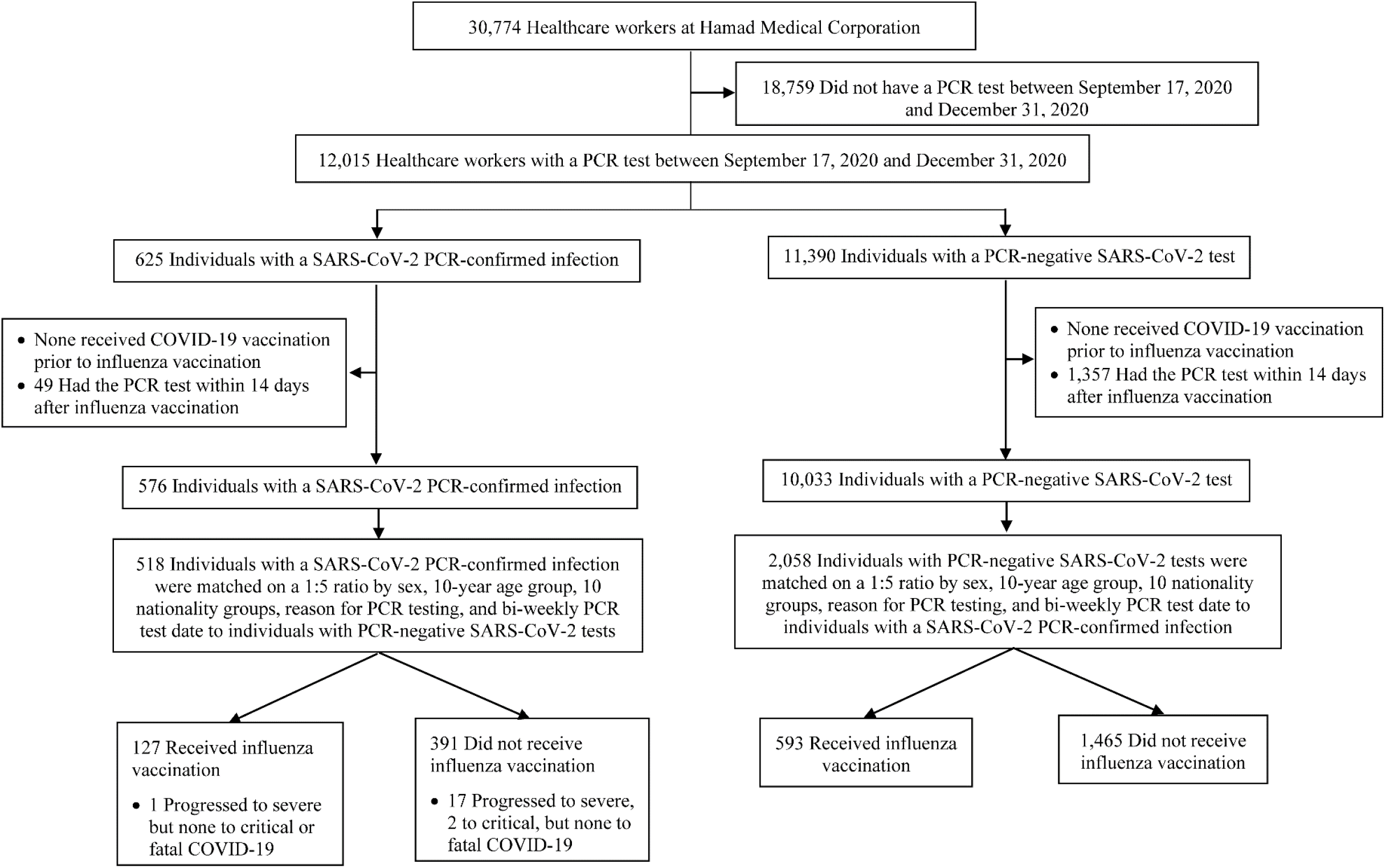
Flowchart describing the population selection process for investigating effectiveness of influenza vaccination against SARS-CoV-2 infection.

Table 1 describes the demographic characteristics of cases and controls. The median age was 36 years (interquartile range (IQR), 32-44) for cases and 35 years (IQR, 32-42) for controls in the matched samples. Slightly less than half (∼ 46%) of cases and controls were males. Participants were of diverse nationalities, and nearly two-thirds were tested because of COVID-like symptoms. Matched study groups were well balanced across the matching factors. The median duration between influenza vaccination and the PCR test was 43 days (IQR, 29-62).

**Table 1.** Demographic characteristics of cases (healthcare workers with PCR-positive tests) and controls (healthcare workers with PCR-negative tests).

| Characteristics | All cases and controls (unmatched) |  |  | Matched cases and controls* |  |  |
| --- | --- | --- | --- | --- | --- | --- |
|  | Cases<br>(PCR-confirmed infection) | Controls<br>(PCR-negative) | SMD† | Cases<br>(PCR-confirmed infection) | Controls<br>(PCR-negative) | SMD† |
| **No** | 576 | 10,033 |  | 518 | 2,058 |  |
| **Median age (IQR) — years** | 37 (32-45) | 36 (32-44) | 0.04 | 36 (32-44) | 35 (32-42) | 0.11 |
| **Age group — no. (%)** |  |  |  |  |  |  |
| <30 years | 80 (13.9) | 1,370 (13.7) | 0.13 | 65 (12.6) | 233 (11.3) | 0.18 |
| 30-39 years | 261 (45.3) | 4,973 (49.6) |  | 255 (49.2) | 1,152 (56.0) |  |
| 40-49 years | 145 (25.2) | 2,354 (23.5) |  | 128 (24.7) | 496 (24.1) |  |
| 50-59 years | 81 (14.1) | 1,120 (11.2) |  | 66 (12.7) | 171 (8.3) |  |
| 60-69 years | 9 (1.6) | 201 (2.0) |  | 4 (0.8) | 6 (0.3) |  |
| 70+ years | 0 (0.0) | 15 (0.2) |  | -- | -- |  |
| **Sex** |  |  |  |  |  |  |
| Male | 270 (46.9) | 4,949 (49.3) | 0.05 | 243 (46.9) | 951 (46.2) | 0.01 |
| Female | 306 (53.1) | 5,084 (50.7) |  | 275 (53.1) | 1,107 (53.8) |  |
| **Nationality‡** |  |  |  |  |  |  |
| Bangladeshi | 1 (0.2) | 77 (0.8) | 0.15 | -- | -- | 0.15 |
| Egyptian | 30 (5.2) | 510 (5.1) |  | 21 (4.1) | 62 (3.0) |  |
| Filipino | 78 (13.5) | 1,637 (16.3) |  | 72 (13.9) | 271 (13.2) |  |
| Indian | 196 (34.0) | 3,095 (30.9) |  | 185 (35.7) | 814 (39.6) |  |
| Nepalese | 6 (1.0) | 45 (0.5) |  | 1 (0.2) | 4 (0.2) |  |
| Pakistani | 11 (1.9) | 223 (2.2) |  | 6 (1.2) | 11 (0.5) |  |
| Qatari | 78 (13.5) | 1,412 (14.1) |  | 74 (14.3) | 262 (12.7) |  |
| Sri Lankan | 1 (0.2) | 22 (0.2) |  | -- | -- |  |
| Sudanese | 24 (4.2) | 347 (3.5) |  | 18 (3.5) | 43 (2.1) |  |
| Other nationalities§ | 151 (26.2) | 2,665 (26.6) |  | 141 (27.2) | 591 (28.7) |  |
| **Reason for PCR testing** |  |  |  |  |  |  |
| Clinical suspicion | 320 (55.6) | 3,680 (36.7) | 0.57 | 299 (57.7) | 1,264 (61.4) | 0.14 |
| Contact tracing | 105 (18.2) | 1,197 (11.9) |  | 95 (18.3) | 346 (16.8) |  |
| Other survey testing | 13 (2.3) | 399 (4.0) |  | 10 (1.9) | 21 (1.0) |  |
| Port of entry | 52 (9.0) | 1,359 (13.6) |  | 43 (8.3) | 172 (8.4) |  |
| Individual request | 18 (3.1) | 535 (5.3) |  | 11 (2.1) | 19 (0.9) |  |
| Survey testing | 37 (6.4) | 1,081 (10.8) |  | 32 (6.2) | 128 (6.2) |  |
| Health care routine testing | 30 (5.2) | 1,517 (15.1) |  | 27 (5.2) | 103 (5.0) |  |
| Pre-travel | 1 (0.2) | 265 (2.6) |  | 1 (0.2) | 5 (0.2) |  |

Abbreviations: IQR, interquartile range; PCR, polymerase chain reaction; SMD, standardized mean difference.
\*Cases and controls were exact-matched on a 1:5 ratio by sex, 10-year age group, 10 nationality groups, reason for PCR testing, and bi-weekly PCR test date.
†SMD is the difference in the mean of a covariate between groups divided by the pooled standard deviation.
‡SMD reported here is for the mean difference between groups divided by the pooled standard deviation.
§Nationalities were chosen to represent the most numerous groups in the population of Qatar.
¶There were 22 other nationalities in the total case population, 74 other nationalities in the total control population, 22 other nationalities among matched cases, and 38 other nationalities among matched controls.

The estimated effectiveness of influenza vaccination against SARS-CoV-2 infection >14 days after receiving the vaccine was 29.7% (95% CI: 5.5-47.7%). All sensitivity analyses yielded consistent results confirming main analysis results (Table 2).

**Table 2.** Effectiveness of influenza vaccination against SARS-CoV-2 infection >14 days after vaccine administration and against any severe, critical, or fatal COVID-19.

| Analyses | Cases*<br>(PCR-confirmed infection) |  | Controls*<br>(PCR-negative tests) |  | Adjusted odds ratio<br>(95% CI) | Effectiveness in %<br>(95% CI)<sup>†</sup> |
| --- | --- | --- | --- | --- | --- | --- |
|  | Vaccinated | Unvaccinated | Vaccinated | Unvaccinated |  |  |
| **Main analysis-Effectiveness against SARS-CoV-2 infection** | 127 | 391 | 593 | 1,465 | 0.70<br>(0.52-0.95) | 29.7<br>(5.5-47.7) |
| **Sensitivity analyses** |  |  |  |  |  |  |
| Adjusting for prior infection in the conditional logistic regression | 127 | 391 | 593 | 1,465 | 0.71<br>(0.53-0.96) | 28.6<br>(4.0-46.9) |
| Excluding prior infection | 126 | 385 | 576 | 1,440 | 0.72<br>(0.54-0.96) | 28.1<br>(3.7-46.3) |
| Excluding travel-related PCR testing | 124 | 357 | 577 | 1,336 | 0.72<br>(0.54-0.97) | 27.9<br>(3.2-46.3) |
| Including all PCR positive and PCR negative tests | 169 | 494 | 844 | 1,917 | 0.66<br>(0.52-0.85) | 33.6<br>(15.0-48.2) |
| **Severity analysis-Effectiveness against any severe, critical, or fatal COVID-19<sup>‡</sup>** | 1 | 19 | 18 | 62 | 0.11<br>(0.01-0.96) | 88.9<br>(4.1-98.7) |

Abbreviations: CI, confidence interval; COVID-19, coronavirus disease 2019; PCR, polymerase chain reaction.
\*Cases and controls were exact-matched on a 1:5 ratio by sex, 10-year age group, 10 nationality groups, reason for PCR testing, and bi-weekly PCR test date.
<sup>†</sup>Vaccine effectiveness was estimated using the test-negative, case-control study design<sup>23,24</sup>.
<sup>‡</sup>Severity<sup>14</sup>, criticality<sup>14</sup>, and fatality<sup>15</sup> were defined as per World Health Organization guidelines.

Of the 127 PCR-positive cases who received influenza vaccination in the matched samples, only 1 progressed to severe, but none to critical or fatal COVID-19 (Figure 1). Meanwhile, of the 391 PCR-positive cases who did not receive influenza vaccination, 17 progressed to severe, 2 to critical, but none to fatal COVID-19. The estimated effectiveness of influenza vaccination against any severe, critical, or fatal COVID-19 was 88.9% (95% CI: 4.1-98.7%; Table 2).

## Discussion

Recent influenza vaccination was associated with a 30% reduction in the risk of SARS-CoV-2 infection by the original virus, before introduction of variants of concern. Recent influenza vaccination was also associated with a 90% reduction in the risk of severe COVID-19, but the 95% confidence interval of the effectiveness estimate was wide. Incidence of severe COVID-19 was rare among those vaccinated, with only one case of severe COVID-19 documented among them.

These results for a HCW population where the influence of the healthy user effect is perhaps minimized, support the conclusion that recent influenza vaccination has a genuine biological effect in protecting against SARS-CoV-2 infection and COVID-19 severity. The findings also corroborate findings of studies that found similar protective effects for influenza vaccination^3-12^, though other studies did not^40,41^. These findings may be explained by influenza vaccination triggering a nonspecific immune activation, or trained or bystander immunity that is protective against SARS-CoV-2 infection^3-5,7,42-47^.

This study has limitations. Effectiveness of only recent influenza vaccination was investigated. The analysis did not factor influenza vaccination from prior seasons. However, given that nonspecific immune activation may not last beyond few weeks, and in context of the relatively rapid waning of COVID-19 vaccine immunity^17,25,48-50^, the observed effectiveness of influenza vaccination is likely short lived. Matching was done for sex, age, nationality, reason for PCR testing, and bi-weekly PCR test date, but it was not possible for other factors, such as comorbidities. However, matching by these factors successfully controlled bias in our earlier studies^17,20,22,25,37^. The majority of HCWs in our sample were also young and less likely to be affected by comorbidities. Nonetheless, one cannot exclude the possibility that in real-world data, bias could arise in unexpected ways, or from unknown sources, such as subtle differences or changes in test-seeking behavior. With the young and occupational nature of our population, these findings may not generalize to the elderly population or to the wider general population.

Notwithstanding these limitations, extensive sensitivity and additional analyses were conducted to investigate effects of potential bias in this study and in our earlier studies that used the same methodology. These included different adjustments and controls in the analysis and different study inclusion and exclusion criteria, to investigate whether effectiveness estimates could have been biased^17,25^. These analyses showed consistent findings^17,25,28,51,52^.

In conclusion, recent influenza vaccination is associated with an appreciable reduction in the risk of SARS-CoV-2 infection and COVID-19 severity. The findings support benefits for influenza vaccination that extend beyond protection against influenza infection and severe disease.

## Data Availability

The dataset of this study is a property of the Qatar Ministry of Public Health that was provided to the researchers through a restricted-access agreement that prevents sharing the dataset with a third party or publicly. Future access to this dataset can be considered through a direct application for data access to Her Excellency the Minister of Public Health (https://www.moph.gov.qa/english/Pages/default.aspx). Aggregate data are available within the manuscript and its Supplementary information.

## Sources of support and acknowledgements

We acknowledge the many dedicated individuals at Hamad Medical Corporation, the Ministry of Public Health, the Primary Health Care Corporation, Qatar Biobank, Sidra Medicine, and Weill Cornell Medicine-Qatar for their diligent efforts and contributions to make this study possible.

The authors are grateful for institutional salary support from the Biomedical Research Program and the Biostatistics, Epidemiology, and Biomathematics Research Core, both at Weill Cornell Medicine-Qatar, as well as for institutional salary support provided by the Ministry of Public Health, Hamad Medical Corporation, and Sidra Medicine. The funders of the study had no role in study design, data collection, data analysis, data interpretation, or writing of the article.

Statements made herein are solely the responsibility of the authors.

## Author contributions

ET, SA, MAA, and AAK co-conceived and co-designed the study, conducted literature review, co-developed the database, and co-wrote the first draft of the article. HC co-designed the study, co-developed the database, performed the statistical analyses, and co-wrote the first draft of the article. LJA co-designed the study, led the statistical analyses, and co-wrote the first draft of the article. All authors contributed to data collection and acquisition, database development, discussion and interpretation of the results, and to the writing of the manuscript. All authors have read and approved the final manuscript.

## Supplementary Appendix

### Supplementary Section S1. COVID-19 severity, criticality, and fatality classification

Severe Coronavirus Disease 2019 (COVID-19) disease was defined per the World health Organization (WHO) classification as a severe acute respiratory syndrome coronavirus 2 (SARS-CoV-2) infected person with “oxygen saturation of <90% on room air, and/or respiratory rate of >30 breaths/minute in adults and children >5 years old (or ≥60 breaths/minute in children <2 months old or ≥50 breaths/minute in children 2–11 months old or ≥40 breaths/minute in children 1–5 years old), and/or signs of severe respiratory distress (accessory muscle use and inability to complete full sentences, and, in children, very severe chest wall indrawing, grunting, central cyanosis, or presence of any other general danger signs)”^1^. Detailed WHO criteria for classifying SARS-CoV-2 infection severity can be found in the WHO technical report^1^.

Critical COVID-19 disease was defined per WHO classification as a SARS-CoV-2 infected person with “acute respiratory distress syndrome, sepsis, septic shock, or other conditions that would normally require the provision of life sustaining therapies such as mechanical ventilation (invasive or non-invasive) or vasopressor therapy”^1^. Detailed WHO criteria for classifying SARS-CoV-2 infection criticality can be found in the WHO technical report^1^.

COVID-19 death was defined per WHO classification as “a death resulting from a clinically compatible illness, in a probable or confirmed COVID-19 case, unless there is a clear alternative cause of death that cannot be related to COVID-19 disease (e.g. trauma). There should be no period of complete recovery from COVID-19 between illness and death. A death due to COVID-19 may not be attributed to another disease (e.g. cancer) and should be counted independently of preexisting conditions that are suspected of triggering a severe course of COVID-19”. Detailed WHO criteria for classifying COVID-19 death can be found in the WHO technical report^2^.

### Supplementary Section S2. Laboratory methods

#### Real-time reverse-transcription polymerase chain reaction testing

Nasopharyngeal and/or oropharyngeal swabs were collected for PCR testing and placed in Universal Transport Medium (UTM). Aliquots of UTM were: extracted on a QIAsymphony platform (QIAGEN, USA) and tested with real-time reverse-transcription PCR (RT-qPCR) using TaqPath™ COVID-19 Combo Kits (Thermo Fisher Scientific, USA) on an ABI 7500 FAST (Thermo Fisher, USA); tested directly on the Cepheid GeneXpert system using the Xpert Xpress SARS-CoV-2 (Cepheid, USA); or loaded directly into a Roche cobas® 6800 system and assayed with a cobas® SARS-CoV-2 Test (Roche, Switzerland). The first assay targets the viral S, N, and ORF1ab gene regions. The second targets the viral N and E-gene regions, and the third targets the ORF1ab and E-gene regions.

All PCR testing was conducted at the Hamad Medical Corporation Central Laboratory or Sidra Medicine Laboratory, following standardized protocols.

**Table S1.** Strengthening the Reporting of Observational Studies in Epidemiology (STROBE) checklist for case-control studies.

|  | Item No | Recommendation | Main text page |
| --- | --- | --- | --- |
| Title and abstract | 1 | (a) Indicate the study’s design with a commonly used term in the title or the abstract | Abstract |
|  |  | (b) Provide in the abstract an informative and balanced summary of what was done and what was found | Abstract |
| Introduction |  |  |  |
| Background/rationale | 2 | Explain the scientific background and rationale for the investigation being reported | Introduction |
| Objectives | 3 | State specific objectives, including any prespecified hypotheses | Introduction |
| Methods |  |  |  |
| Study design | 4 | Present key elements of study design | Methods (‘Study design’) |
| Setting | 5 | Describe the setting, locations, and relevant dates, including periods of recruitment, exposure, follow-up, and data collection | Methods (‘Study design’) & Figure 1 |
| Participants | 6 | (a) Give the eligibility criteria, and the sources and methods of case ascertainment and control selection. Give the rationale for the choice of cases and controls | Methods (‘Study design’) & Figure 1 |
|  |  | (b) For matched studies, give matching criteria and the number of controls per case |  |
| Variables | 7 | Clearly define all outcomes, exposures, predictors, potential confounders, and effect modifiers. Give diagnostic criteria, if applicable | Methods (‘Study design’), Table 1, & Supplementary Sections S1 and S2 |
| Data sources/measurement | 8 | For each variable of interest, give sources of data and details of methods of assessment (measurement). Describe comparability of assessment methods if there is more than one group | Methods (‘Study design’ & ‘Statistical analysis’), & Supplementary Sections S1 and S2 |
| Bias | 9 | Describe any efforts to address potential sources of bias | Methods (‘Study design’, paragraphs 5-6 & ‘Statistical analysis’) |
| Study size | 10 | Explain how the study size was arrived at | Figure 1 |
| Quantitative variables | 11 | Explain how quantitative variables were handled in the analyses. If applicable, describe which groupings were chosen and why | Methods (‘Study design’), Table 1, & Supplementary Sections S1 and S2 |
| Statistical methods | 12 | (a) Describe all statistical methods, including those used to control for confounding | Methods (‘Statistical analysis’) |
|  |  | (b) Describe any methods used to examine subgroups and interactions | Methods (‘Statistical analysis’) |
|  |  | (c) Explain how missing data were addressed | Not applicable, see Methods (‘Study design’, paragraph 1) |
|  |  | (d) If applicable, explain how matching of cases and controls was addressed | Methods (‘Study design’, paragraph 5 & ‘Statistical analysis’, paragraph 1) |
|  |  | (e) Describe any sensitivity analyses | Methods (‘Statistical analysis’, paragraph 2) |
| Results |  |  |  |
| Participants | 13 | (a) Report numbers of individuals at each stage of study—eg numbers potentially eligible, examined for eligibility, confirmed eligible, included in the study, completing follow-up, and analysed | Results, paragraph 1 & Figure 1 |
|  |  | (b) Give reasons for non-participation at each stage |  |
|  |  | (c) Consider use of a flow diagram |  |
| Descriptive data | 14 | (a) Give characteristics of study participants (eg demographic, clinical, social) and information on exposures and potential confounders | Results, paragraph 2 & Table 1 |
|  |  | (b) Indicate number of participants with missing data for each variable of interest | Not applicable, see Methods (‘Study design’, paragraph 1) |
| Outcome data | 15 | Report numbers in each exposure category, or summary measures of exposure | Results, paragraphs 3-4 & Table 2 |
| Main results | 16 | (a) Give unadjusted estimates and, if applicable, confounder-adjusted estimates and their precision (eg, 95% confidence interval). Make clear which confounders were adjusted for and why they were included | Results, paragraphs 3-4 & Table 2 |
|  |  | (b) Report category boundaries when continuous variables were categorized | Table 1 |
|  |  | (c) If relevant, consider translating estimates of relative risk into absolute risk for a meaningful time period | Not applicable |
| Other analyses | 17 | Report other analyses done—eg analyses of subgroups and interactions, and sensitivity analyses | Results, paragraph 3 & Table 2 |
| Discussion |  |  |  |
| Key results | 18 | Summarise key results with reference to study objectives | Discussion, paragraphs 1-2 |
| Limitations | 19 | Discuss limitations of the study, taking into account sources of potential bias or imprecision. Discuss both direction and magnitude of any potential bias | Discussion, paragraphs 3-4 |
| Interpretation | 20 | Give a cautious overall interpretation of results considering objectives, limitations, multiplicity of analyses, results from similar studies, and other relevant evidence | Discussion, paragraph 5 |
| Generalisability | 21 | Discuss the generalisability (external validity) of the study results | Discussion, paragraph 3 |
| **Other information** |  |  |  |
| Funding | 22 | Give the source of funding and the role of the funders for the present study and, if applicable, for the original study on which the present article is based | Sources of support and acknowledgements |

